# Force Sensor Reduces Measurement Error during Sit-to-Stand Assessment of Cerebral Autoregulation

**DOI:** 10.1101/2022.09.21.22280201

**Authors:** Alicen A. Whitaker, Eric D. Vidoni, Robert N. Montgomery, Kailee Carter, Katelyn Struckle, Sandra A. Billinger

**Author notes:** **Corresponding Author:** Sandra A. Billinger, PT, PhD; Address: 3901 Rainbow Blvd, Mail Stop 3051, Kansas City, KS 66160.

## Abstract

**Introduction:** Novel implementation of a force sensor during a sit-to-stand measure of dynamic cerebral autoregulation (dCA) has been shown to measure the exact moment an individual stands up from a chair, called arise-and-off (AO). Traditional measures estimate time delay (TD) before the onset of the dCA response from the verbal command to stand. We hypothesized that using a force sensor to measure AO would significantly improve the accuracy of the TD measure compared to estimating from verbal command.

**Methods:** Middle cerebral artery blood velocity (MCAv) and mean arterial pressure (MAP) were measured simultaneously during three sit-to-stand measures of dCA. Participants were seated for 60 seconds, then performed a sit-to-stand and the force sensor detected AO. TD was calculated as the time from AO until an increase in cerebrovascular conductance (CVC = MCAv/MAP). TD was also calculated from verbal command to stand.

**Results:** Sixty-five participants completed the study: twenty-five young adults (age 25±2 years), twenty older adults (age 61±13 years), and twenty individuals with stroke (age 60±13 years). There was a significant difference in TD when using AO compared to estimating (F-value=49.9, p<0.001). Estimated TD introduced ∼17% measurement error. Average TD measurement error was not related to age (r =-0.04, p=0.76) or history of stroke (r=0.01, p=0.96).

**Discussion:** The addition of a force sensor to detect AO during a sit-to-stand procedure showed a significant difference in the TD dCA measurement. Our data support the implementation of a force sensor during sit-to-stand dCA measures in healthy adults across all ages and after stroke.

## Introduction

Dynamic cerebral autoregulation (dCA) is the ability of the brain to independently react to changes in peripheral mean arterial pressure (MAP) and maintain cerebrovascular stability.^1-3^ The dCA response can be measured during a common physiological stressor such as a sit-to-stand.^4^ During a sit-to-stand, MAP decreases due to gravity and peripheral vasodilation.Therefore, the dCA response must quickly increase the cerebrovascular conductance (CVC) of middle cerebral artery blood velocity (MCAv) to the brain to maintain homeostasis.^4,5^

One metric of the sit-to-stand dCA response is the time delay (TD) before the onset of the regulation response, or how quickly CVC increases after the sit-to-stand.^4-6^ The TD before the onset of the regulation response typically occurs within ∼10 seconds of standing^4-7^ but can occur on average ∼1.5 seconds after standing in healthy young men.^5^ Therefore, the TD before the onset of the regulation response is an important temporal measure.

While previous studies have shown the ability to implement an accelerometer to detect the angle and speed of the sit-to-stand, ^8-14^ we are the first to implement a custom force sensor to improve the temporal accuracy of the TD response.^15^ Our methodology uses a force sensor to simultaneously measures the sit-to-stand reaction time with the physiological dCA response.^15^ We have reported on the ability to implement a force sensor to identify a defined moment of transition between sitting and standing, or arise-and-off (AO).^15^ Traditionally, the TD response is calculated from the time in which individuals are verbally told to stand up from a seated position. However, in our prior work, we describe the methodology to accurately identify the exact moment of transition during a sit-to-stand recording and show how AO may potentially differ in older adults and individuals post-stroke.^15^

Therefore, the objective of this study was to determine whether implementing a force sensor to detect the exact moment of AO would significantly improve the accuracy of the TD response compared to the traditional estimation of TD from the verbal command to stand. We hypothesized that using a force sensor to measure AO to calculate the TD response during a sit-to-stand measure of dCA would be significantly more accurate than the estimated TD response from the verbal command to stand in healthy young adults, older adults, and individuals with stroke. We also hypothesized that the measurement error of the TD response would be significantly associated with participant demographics such as age, BMI, the ability to stand quickly, or history of stroke.

## Methods

This study reports on the accuracy of current methodology used in an ongoing study (NCT04673994). The inclusion and exclusion criteria of this study have been published previously.^15^ Briefly, we enrolled healthy young adults 18-30 years old with low cardiovascular risk.^16^ Older adults and individuals post-stroke (6 months – 5 years ago), were 1) age 40-80 years old, 2) sedentary (<150 minutes brisk exercise/week),^16^ 3) able to answer consenting questions and follow a 2-step command, 4) able to stand up from a chair without physical assistance, and 5) not diagnosed with another underlying neurological disease. Individuals with chronic stroke were included within this study as they are a clinical population that presents with hemiplegia and a slower sit-to-stand response that may be detected by the force sensor.

The Human Subjects Committee within the University of Kansas Medical Center’s Institutional Review Board approved the study. Prior to starting the study, all individuals were informed of study procedures, benefits, and risks, and asked to provide voluntary written consent. We then collected demographic information.

Our laboratory was kept at a constant temperature (22°C to 24°C) and dimly lit during our recordings. On the day of the visit, participants were asked to take their medications as prescribed, not to have caffeine for 8 hours,^17-19^ not to perform vigorous exercise for 24 hours,^20^ and to abstain from alcohol for 24 hours.^21^ Participants were seated with their feet flat on the ground and an upright trunk posture. To determine whether individuals had the leg strength and balance to perform a sit-to-stand independently, a standardized 5x sit-to-stand was performed. Participants were asked to stand up and sit down 5 times as quickly as they could without the use of their arms and the time it took to perform the 5x sit-to-stand was measured in seconds.^22-24^ Equipment was then donned, which included: 1) bilateral TCD probes (2-MHz, Multigon Industries Inc, Yonkers, New York) to measure MCAv, 2) a plethysmograph (Finometer, Finapres Medical Systems, Amsterdam, the Netherlands) was placed on the left middle finger (or upper extremity without spasticity for individuals post-stroke) to measure beat-to-beat MAP, and 3) a 5-lead electrocardiogram (ECG; Cardiocard, Nasiff Associates, Central Square, New York) to measure heart rate. As we have done previously,^15^ participants were instructed to place their hand with the finger plethysmograph flat on their chest at heart level and were fitted with an arm sling to hold the Finometer in place. ^4,5,25^

Our custom force sensor was then placed underneath the participant at the level of their right ischial tuberosity. For individuals with stroke, the force sensor was placed underneath the non-affected lower extremity.^26^ Participants performed seated rest for 60 seconds. The participant was then given a 3 second countdown and asked to stand at the 60 second mark. The participant continued standing for an additional 2 minutes for hemodynamic stability.^27^ All measures were recorded at 500 Hz using a custom written software within MATLab, implementing the Data Acquisition Toolbox (R2019a, TheMathworks Inc, Natick, Massachusetts). Participants performed 3 sit-to-stand procedures (T1, T2, and T3) during the study visit, each separated by 20 minutes.

### Data Analysis

Offline processing of the collected data was done using custom written software within MATLab. The ECG QRS complex was used to calculate beat-to-beat mean MCAv and MAP. As previously published, AO was identified as the minimum of the second derivative of the recorded force sensor voltage upon standing.^15^ The manual identification of TD before the onset of the regulation response was completed by 2 trained researchers and evaluated as the physiological beat after standing where there was a continuous increase in CVC (CVC = MCAv/MAP).^5,15,28^

The primary aim was to determine whether the force sensor significantly altered the TD response when calculated using AO compared to the estimated time of stance (from 60 seconds). To analyze this aim, the TD response calculated from AO and TD calculated from 60 seconds was compared using with a Two-Way Repeated Measures ANOVA with a within-subjects effect for time (T1, T2, and T3) and type of calculation (using AO versus Estimated). At each time point, post-hoc Wilcoxin Signed Rank t-tests were used to analyze the difference in the TD response calculated from AO or estimated from 60 seconds (verbal command to stand). We also performed a Mixed Model ANOVA to examine differences in the AO response with a within-subjects effect for time (T1, T2, and T3) and between-subjects effect for group (young adult, older adult, and individuals post-stroke).

The force sensor measures the true time of stance with AO, therefore, we wanted to calculate the measurement error between the two methodologies. To analyze measurement error, we calculated the difference between the AO TD response and the estimated TD response from 60 seconds. The average TD measurement error was then plotted within a histogram graph to show the frequency distribution. Spearman correlations were then used to determine if measurement error was significantly related to age, BMI, or the 5x sit-to-stand. A linear regression was used to determine if measurement error was significantly related to whether a participant had a history of stroke. Another way we examined the AO response of each group was to calculate the coefficient of variation (CoV = (standard deviation/mean) * 100).

## Results

In total, 65 individuals including healthy young adults (n = 25), older adults (n = 20), and individuals with stroke (n = 20), completed the sit-to-stand transition with the force sensor. Due to noise in MAP and TCD with standing, 59 individuals had complete data sets with a TD response at all three timepoints. Participant characteristics are shown in **Table 1**.

**Table 1.**
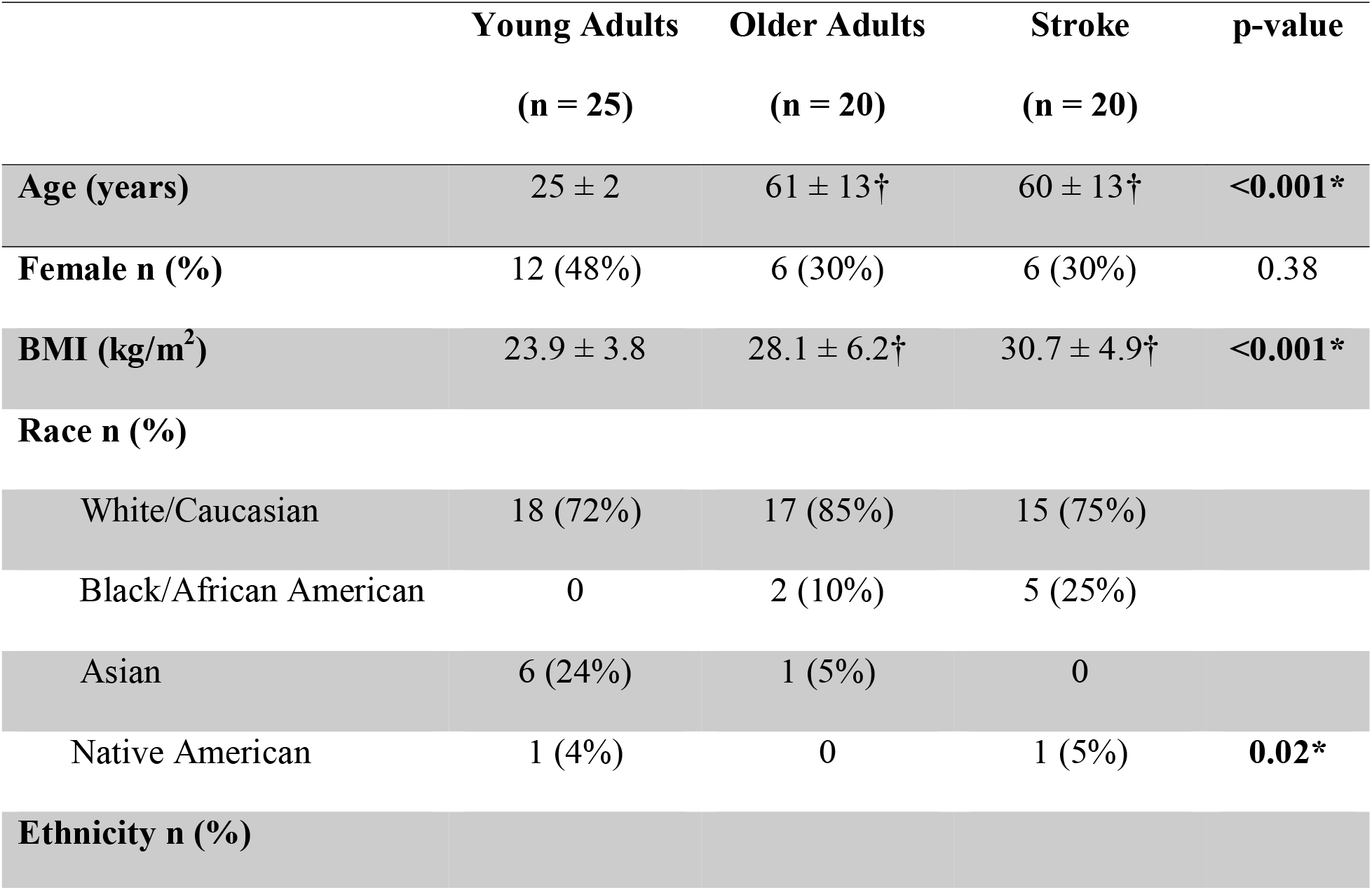
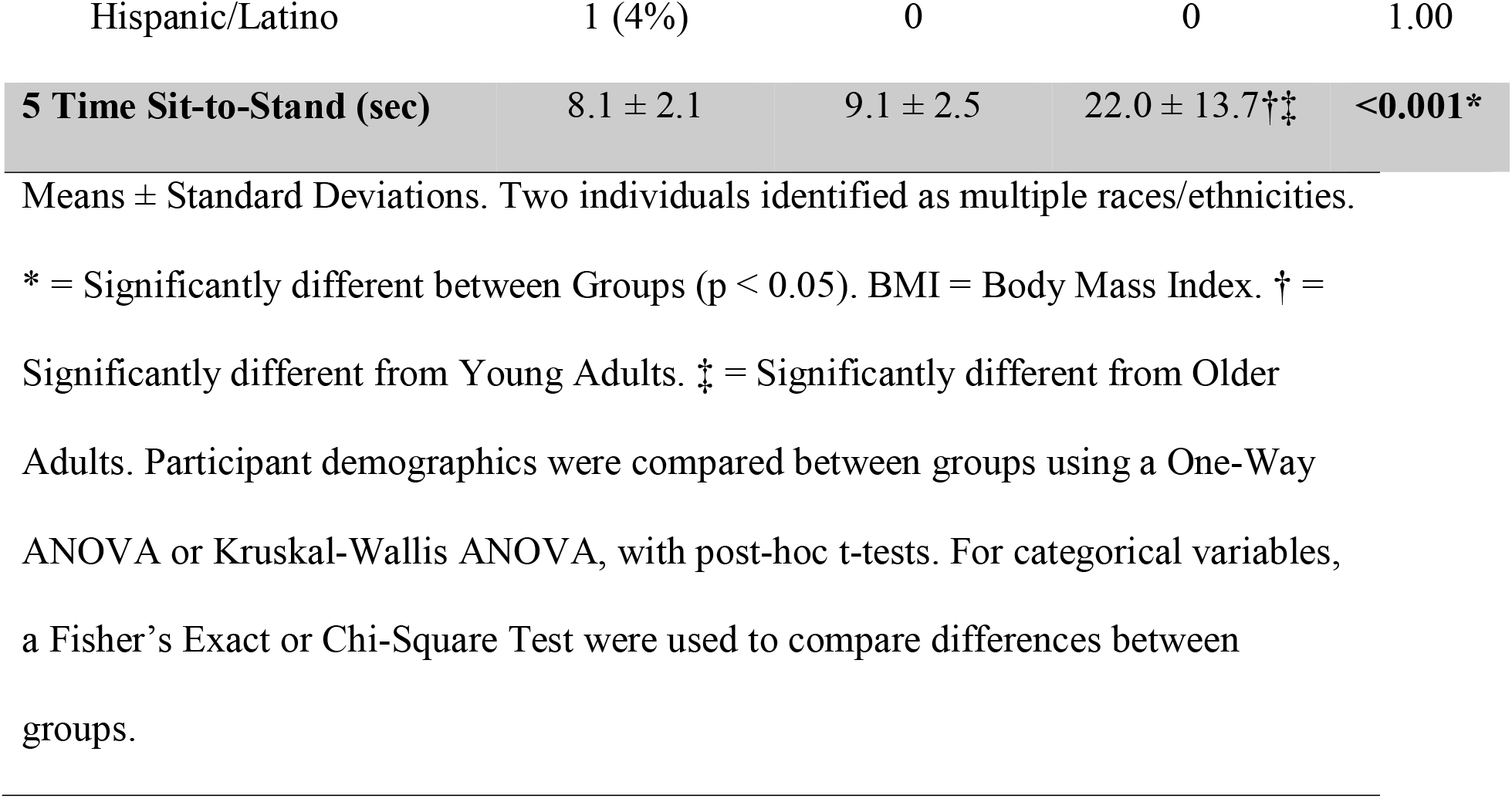
Participant Characteristics.

### Comparing the TD Response using AO vs. Estimating

Our primary analysis was to determine whether the force sensor significantly improved the accuracy of the TD response during a sit-to-stand measure of dCA compared to estimating TD from when participants were asked to stand at 60 seconds into the recording. Our primary analysis showed a significant difference in TD when using AO compared to estimating from the verbal command to stand at 60 seconds (F-value = 49.9, p < 0.001). Across the 3 trials within the study visit, we found that the TD response was not significantly different across the three time points (p = 0.36), which suggests potential stability in a same day measure of the TD response. When performing post-hoc comparisons, the TD response at each separate time point was significantly shorter when using AO compared to estimating from 60 seconds (p ≤ 0.001). At T1, T2, and T3, the respective AO TD response was 3.2 ± 2.3 seconds, 2.8 ± 2.0 seconds, and 2.9 ± 2.2 seconds compared to the estimated TD response of 3.6 ± 2.4 seconds, 3.2 ± 2.0 seconds, and 3.2 ± 2.3 seconds. We also wanted to determine whether AO significantly differed between young adults, older adults, and individuals post-stroke. We found that the AO response was not significantly different between groups (p = 0.85) or across the different time points (p = 0.63).

### Measurement Error

We have previously published on the ability to mark the true time of stance (AO) during a sit-to-stand recording of the dCA response.^15^ By calculating the measurement error, we were able to examine the error in the calculation of the estimated TD response from the time in which individuals were asked to stand (60 seconds). A histogram reveals the frequency of how predicted values differ from the true value, shown in **Figure 1**. The distribution of measurement error is negatively skewed showing that most individuals stood up ∼0.5 seconds after being told to stand. With the average TD response being ∼3 seconds, estimating the TD response from the verbal command to stand would introduce ∼17% measurement error.

**Figure 1.**
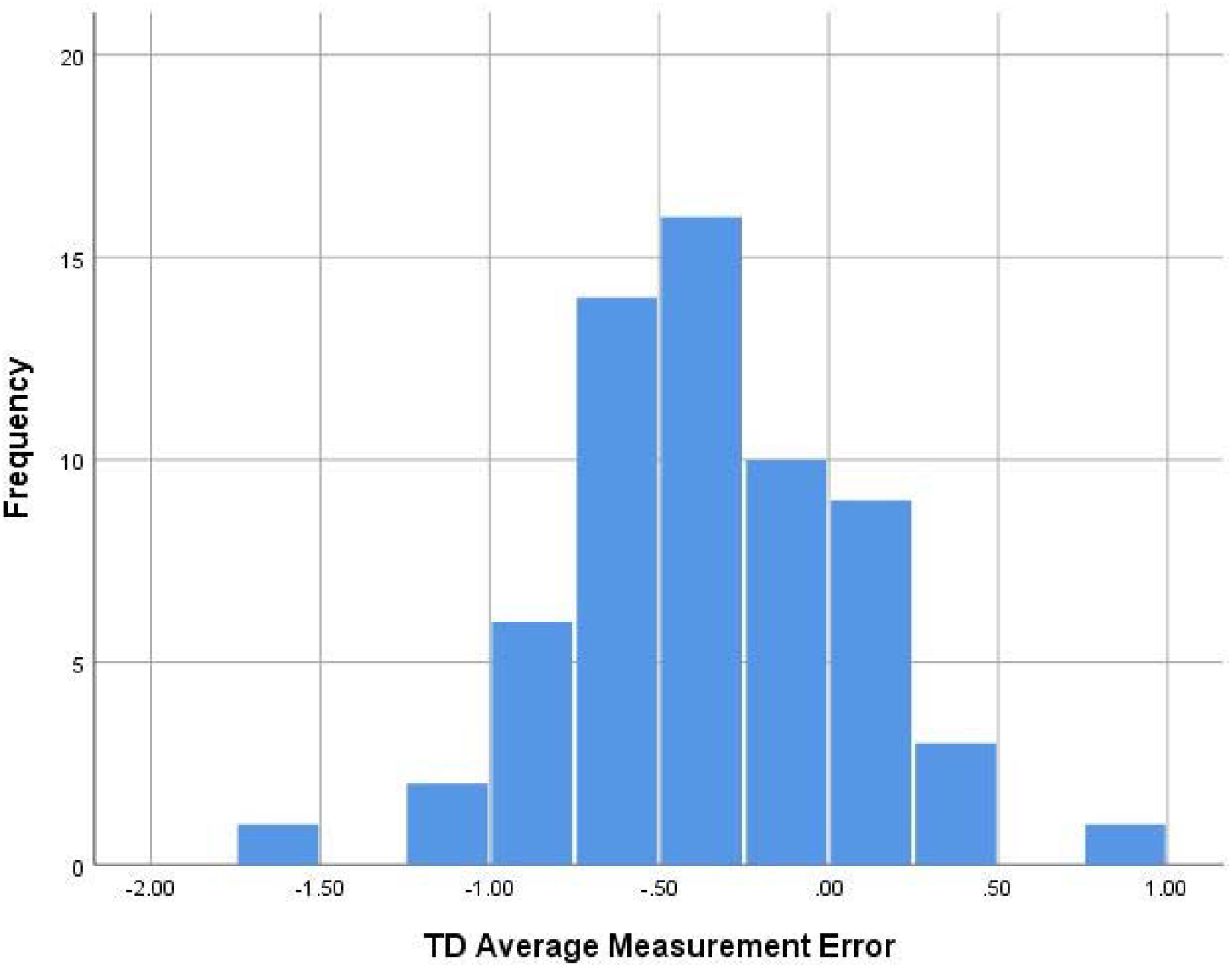
Histogram of the TD Average Measurement Error. TD = Time Delay.

The average measurement error of the TD response was not significantly related to age (r = -0.04, p = 0.76), BMI (r = -0.05, p = 0.71), 5x sit-to-stand (r = -0.07, p = 0.58), or history of stroke (r = 0.04, p = 0.76).

### Variation in AO Between Groups

Though the measurement error was not significantly related to age or history of stroke, we calculated the CoV in AO to reveal any potential variations in the AO response between groups and over time, shown in **Table 2**. The CoV of AO is small in all groups, showing stable methodology for all individuals. Examining the values of the CoV shows low variability across timepoints and groups.

**Table 2.** Coefficient of Variation of AO.

|  | **T1** | **T2** | **T3** |
| --- | --- | --- | --- |
|  | **AO CoV** | **AO CoV** | **AO CoV** |
| **Young Adults** | 0.89% | 0.79% | 0.84% |
| **Older Adults** | 0.73% | 1.22% | 0.78% |
| **Stroke** | 1.11% | 0.86% | 0.72% |

## Discussion

Our findings in a diverse group of participants support our hypothesis and show that using a force sensor to detect AO during a sit-to-stand measure of dCA shows a significant improvement in the accuracy of the TD response calculation compared to the traditional method of estimating TD from the time in which individuals were asked to stand. The main findings of this study include 1) the TD response was significantly faster when calculated using AO compared to estimating from 60 seconds, 2) the force sensor reduced measurement error for all individuals, and 3) the AO response showed small variability which does not differ between measures taken within the same day or between healthy young adults, older adults, or individuals post-stroke.

Our methodology of implementing a force sensor has shown that variability in the transition between sitting and standing can potentially reduce measurement error by ∼17% within the TD calculation. The reduction in measurement error ensures more accurate reporting of the transient time course changes in MCAv and MAP, which is critical in the field of cerebrovascular physiology. Further, there is considerable interest in studying older adults and clinical populations such as traumatic brain injury, Alzheimer’s disease and dementia, and stroke, where the nature of these conditions may introduce inherent variability. Therefore, refinement of the sit-to-stand methodology with the use of the force sensor could have a significant impact on data interpretation, reporting of findings and allowing the field to address important gaps in knowledge regarding cerebral autoregulation in humans. This is supported by our data that the reduction in measurement error was significant for all individuals regardless of age or clinical diagnosis of stroke. Our data presented here show no significant difference across the three time points within a single session.

While prior studies have implemented an accelerometer to normalize the dCA response to the speed of the sit-to-stand, the accelerometer was not temporally aligned with the physiological dCA response and the moment of stance was not determined.^8-13^ Implementation of a force sensor will address this limitation by measuring the exact time of stance (AO) during the dCA measure. Therefore, implementing both the force sensor and accelerometer may improve the accuracy of sit-to-stand dCA measures by accounting for both the timing and speed of the sit-to-stand. While our intention was to employ a quick sit to stand response (0-3 seconds) aligned with the literature,^5^ our initial work showed that the stance time in older adults and individuals with cerebrovascular disease may be altered.^15^ Future studies are needed to report on the simultaneous use of a force sensor and accelerometer to increase the accuracy of the sit-to-stand dCA response.

In conclusion, we compared whether a force sensor applied to the sit-to-stand procedure would reduce measurement error in a diverse group of individuals. The force sensor appears to significantly decrease measurement error in humans with low variability across time points in a single session.

## Data Availability

All data are available upon request.

## References

1. Roy CS, Sherrington CS. On the Regulation of the Blood-supply of the Brain. J Physiol. 1890;11(1-2):85–158 117.

2. Brassard P, Labrecque L, Smirl JD, et al. Losing the dogmatic view of cerebral autoregulation. Physiol Rep. 2021;9(15):e14982.

3. van Beek AH, Claassen JA, Rikkert MG, Jansen RW. Cerebral autoregulation: an overview of current concepts and methodology with special focus on the elderly. J Cereb Blood Flow Metab. 2008;28(6):1071–1085.

4. Sorond FA, Serrador JM, Jones RN, Shaffer ML, Lipsitz LA. The sit-to-stand technique for the measurement of dynamic cerebral autoregulation. Ultrasound Med Biol. 2009;35(1):21–29.

5. Labrecque L, Rahimaly K, Imhoff S, et al. Diminished dynamic cerebral autoregulatory capacity with forced oscillations in mean arterial pressure with elevated cardiorespiratory fitness. Physiol Rep. 2017;5(21).

6. Labrecque L, Rahimaly K, Imhoff S, et al. Dynamic cerebral autoregulation is attenuated in young fit women. Physiol Rep. 2019;7(2):e13984.

7. Serrador JM, Sorond FA, Vyas M, Gagnon M, Iloputaife ID, Lipsitz LA. Cerebral pressure-flow relations in hypertensive elderly humans: transfer gain in different frequency domains. J Appl Physiol (1985). 2005;98(1):151–159.

8. Barnes SC, Ball N, Haunton VJ, Robinson TG, Panerai RB. The cerebrocardiovascular response to periodic squat-stand maneuvers in healthy subjects: a time-domain analysis. Am J Physiol Heart Circ Physiol. 2017;313(6):H1240–H1248.

9. Barnes SC, Ball N, Panerai RB, Robinson TG, Haunton VJ. Random squat/stand maneuvers: a novel approach for assessment of dynamic cerebral autoregulation? J Appl Physiol (1985). 2017;123(3):558–566.

10. Barnes SC, Ball N, Haunton VJ, Robinson TG, Panerai RB. How many squat-stand manoeuvres to assess dynamic cerebral autoregulation? Eur J Appl Physiol. 2018;118(11):2377–2384.

11. Barnes SC, Haunton VJ, Beishon L, Llwyd O, Robinson TG, Panerai RB. Extremes of cerebral blood flow during hypercapnic squat-stand maneuvers. Physiol Rep. 2021;9(19):e15021.

12. Batterham AP, Panerai RB, Robinson TG, Haunton VJ. Does depth of squat-stand maneuver affect estimates of dynamic cerebral autoregulation? Physiol Rep. 2020;8(16):e14549.

13. Panerai RB, Batterham A, Robinson TG, Haunton VJ. Determinants of cerebral blood flow velocity change during squat-stand maneuvers. Am J Physiol Regul Integr Comp Physiol. 2021;320(4):R452–R466.

14. Klein T, Bailey TG, Wollseiffen P, Schneider S, Askew CD. The effect of age on cerebral blood flow responses during repeated and sustained stand to sit transitions. Physiol Rep. 2020;8(9):e14421.

15. Whitaker AA, Vidoni ED, Aaron SE, Rouse AG, Billinger SA. Novel application of a force sensor during sit-to-stands to measure dynamic cerebral autoregulation onset. Physiol Rep. 2022;10(7):e15244.

16. Thompson PD, Arena R, Riebe D, Pescatello LS, American College of Sports M. ACSM’s new preparticipation health screening recommendations from ACSM’s guidelines for exercise testing and prescription, ninth edition. Curr Sports Med Rep. 2013;12(4):215–217.

17. Perod AL, Roberts AE, McKinney WM. Caffeine can affect velocity in the middle cerebral artery during hyperventilation, hypoventilation, and thinking: a transcranial Doppler study. J Neuroimaging. 2000;10(1):33–38.

18. In: Caffeine for the Sustainment of Mental Task Performance: Formulations for Military Operations. Washington (DC)2001.

19. Addicott MA, Yang LL, Peiffer AM, et al. The effect of daily caffeine use on cerebral blood flow: How much caffeine can we tolerate? Hum Brain Mapp. 2009;30(10):3102–3114.

20. Burma JS, Copeland P, Macaulay A, Khatra O, Wright AD, Smirl JD. Dynamic cerebral autoregulation across the cardiac cycle during 8 hr of recovery from acute exercise. Physiol Rep. 2020;8(5):e14367.

21. Mathew RJ, Wilson WH. Regional cerebral blood flow changes associated with ethanol intoxication. Stroke. 1986;17(6):1156–1159.

22. Mong Y, Teo TW, Ng SS. 5-repetition sit-to-stand test in subjects with chronic stroke: reliability and validity. Arch Phys Med Rehabil. 2010;91(3):407–413.

23. Tiedemann A, Shimada H, Sherrington C, Murray S, Lord S. The comparative ability of eight functional mobility tests for predicting falls in community-dwelling older people. Age Ageing. 2008;37(4):430–435.

24. Ng S. Balance ability, not muscle strength and exercise endurance, determines the performance of hemiparetic subjects on the timed-sit-to-stand test. Am J Phys Med Rehabil. 2010;89(6):497–504.

25. Lipsitz LA, Mukai S, Hamner J, Gagnon M, Babikian V. Dynamic regulation of middle cerebral artery blood flow velocity in aging and hypertension. Stroke. 2000;31(8):1897–1903.

26. Brunt D, Greenberg B, Wankadia S, Trimble MA, Shechtman O. The effect of foot placement on sit to stand in healthy young subjects and patients with hemiplegia. Arch Phys Med Rehabil. 2002;83(7):924–929.

27. Drapeau A, Labrecque L, Imhoff S, et al. Six weeks of high-intensity interval training to exhaustion attenuates dynamic cerebral autoregulation without influencing resting cerebral blood velocity in young fit men. Physiol Rep. 2019;7(15):e14185.

28. Lind-Holst M, Cotter JD, Helge JW, et al. Cerebral autoregulation dynamics in endurance-trained individuals. J Appl Physiol (1985). 2011;110(5):1327–1333.

